# FORECASTING COVID-19 PANDEMIC: A DATA-DRIVEN ANALYSIS

**DOI:** 10.1101/2020.05.12.20099192

**Authors:** Khondoker Nazmoon Nabi

## Abstract

In this paper, a new Susceptible-Exposed-Symptomatic Infectious-Asymptomatic Infectious-Quarantined-Hospitalized-Recovered-Dead (*SEI_D_I_U_QHRD*) deterministic compartmental model has been proposed and calibrated for interpreting the transmission dynamics of the novel coronavirus disease (COVID-19). The purpose of this study is to give a tentative prediction of the epidemic peak for Russia, Brazil, India and Bangladesh which could become the next COVID-19 hotspots in no time by using a Trust-region-reflective (TRR) algorithm which one of the well-known real data fitting techniques. Based on the publicly available epidemiological data from late January until 10 May, it has been estimated that the number of daily new symptomatic infectious cases for the above mentioned countries could reach the peak around the beginning of June with the peak size of ~ 15, 774 (95% CI, 13,814-17,734) symptomatic infectious cases in Russia, ~ 26, 449 (95% CI, 23,489-29,409) cases in Brazil, ~ 9, 504 (95% CI, 8,378-10,630) cases in India and ~ 2, 209 (95% CI, 1,878-2,540) cases in Bangladesh. As of May 11, 2020, incorporating the infectiousness capability of asymptomatic carriers, our analysis estimates the value of the basic reproduction number (*R*_0_) as of May 11, 2020 was found to be ~ 4.234 (95% CI, 3.764-4.7) in Russia,~ 5.347 (95% CI, 4.737-5.95) in Brazil, ~ 5.218 (95% CI, 4.56-5.81)in India, ~ 4.649 (95% CI, 4.17-5.12) in the United Kingdom and ~ 3.53 (95% CI, 3.12-3.94) in Bangladesh. Moreover, Latin hypercube sampling-partial rank correlation coefficient (LHS-PRCC) which is a global sensitivity analysis (GSA) method is applied to quantify the uncertainty of our model mechanisms, which elucidates that for Russia, the recovery rate of undetected asymptomatic carriers, the rate of getting home-quarantined or self-quarantined and the transition rate from quarantined class to susceptible class are the most influential parameters, whereas the rate of getting home-quarantined or self-quarantined and the inverse of the COVID-19 incubation period are highly sensitive parameters in Brazil, India, Bangladesh and the United Kingdom which could significantly affect the transmission dynamics of the novel coronavirus. Our analysis also suggests that relaxing social distancing restrictions too quickly could exacerbate the epidemic outbreak in the above-mentioned countries.

## 1. Introduction

The coronavirus disease 2019 (COVID-19) has evolved as a global public health emergency affecting 212 countries and territories around the world as of May 12, 2020 Worldometer, (2020). COVID-19 is one kind of respiratory disease by the novel coronavirus (SARS-CoV-2) that was first spotted around late December 2019, in Wuhan, Hubei province, China Li et al., (2020); Zhou et al., (2020). This novel virus started transmitting around the world rapidly, and on 30 January, WHO declared the outbreak as a Public Health Emergency of International Concern (PHEIC). Later, WHO Director-General announced COVID-19 as a global pandemic. As of May 11, 2020, an outbreak of COVID-19 has resulted in 4,271,689 confirmed cumulative cases with reported deaths of 287,613 worldwide Worldometer, (2020). In most of the countries, infected patients are struggling to get the proper treatment due to highly transmissible and virulent nature of the virus. Nevertheless, numerous COVID-19 mitigation strategies has been adapted so far such as quarantine, isolation, promoting the wearing of face masks, travel restrictions, and lockdowns with a view to reducing community transmission of the disease.

In the absence of either established effective treatment or a vaccine, the already fragile health care systems of different developed and developing countries can be over-burdened due to the overwhelming surge of infections in in the coming months provided that the spread is not controlled. This pandemic is giving an upsurge to multifarious noteworthy socio-economic and public health impacts and has highlighted the significance of detecting the evolution of the disease and prediction of disease future dynamics for designing infectious disease prevention and control strategies, effective public health policies and economical activity guidelines. Different mathematical paradigms have always played a notable role in providing deeper understanding of the transmission mechanisms of a disease outbreak, contributing considerable insights for controlling the disease outbreak. One of the familiar models for human-to-human transmission which is reasonably predictive is Susceptible-Infectious-Removed (SIR) epidemic model proposed by Kermack-Mckendrick in 1927 Calafiore et al., (2020); Nesteruk (2020). After that, the SIR epidemic model has been extended to Susceptible-Exposed-Infectious-Removed (SEIR) model and many of its variants to explore the risk factors of a disease or predict the dynamics of a disease outbreak Wu et al., (2020); Calafiore et al.. (2020); kucharski et al., (2020); Simha et al., (2020); Anastassopoulou et al., (2020); Nesteruk (2020). It is really challenging in population based model to incorporate certain real-world complexities. In fact, analysis and prediction could go wrong in the absence of adequate historical real data. On the other hand, various agent-based often stochastic models where individuals interact on a network structure and get infected stochastically, have been treated as useful tools for tracing fine-grained effects of heterogeneous intervention policies in diverse disease outbreaks Ferguson et al., (2020); Chang et al., (2020); Wilder et al., (2020); Ruiz et al., (2020). However, accuracy of this approach can be a vital issue due to the time-varying nature of network-structure.

Since the outbreak of the virus, incorporating travel between major cities in China, Li et al., (2020) proposed an SEIR-model incorporating a metapopulation structure for both reported and unreported infections. It has been discovered in their studies that around 86% of all cases went undetected in Wuhan before travel restrictions imposed on January 23, 2020. According to their estimation, on an individual basis, around 55% asymptomatic spreaders were contagious who were responsible for 79% of new infected cases. Other studies Ferguson et al., (2020); Verity et al., (2020) solidified the significance of incorporating asymptomatic carriers with a view to understanding COVID-19 future dynamics appropriately. Later, Calafiore et al., (2020) estimated that around 63% cases went under-reported in Italy by analyzing a modified SIR-model. Anastassopoulou et al., (2020) apply the SIRD model to Chinese official statistics, estimating parameters using linear regression and predict the COVID-19 pandemic in Hubei province by using these models and parameters. Nonetheless, long-term forecasting is debatable while using a simple mathematical model. Diego Caccavo, (2020) and Peter Turchin, (2020) independently apply modified SIRD models, in which parameters change overtime following specific function forms. Parameters govern these functions are estimated by minimizing the sum-of-square-error. However, using the sum-of-square method causes over-fitting and always favors a complex model, therefore it is not suitable to access policy effectiveness. Moreover, fitting the SIRD model in the early stage of infection is questionable as well.

In the light of above shortcomings of several established mathematical models, a more refined deterministic system of nonlinear differential equations, considering all possible interactions, a Susceptible-Exposed-Symptomatic Infectious-Asymptomatic Infectious-Quarantined-Hospitalized-Recovered-Dead (*SEI_D_I_U_QHRD*) model has been proposed, which can give more accurate and robust short-term as well as long-term predictions of COVID-19 future dynamics. This model could be considered as a generalization of SEIR, which is based on the introduction of asymptomatic infectious state, quarantined state in order to understand the effect of preventive actions and hospitalized (isolated) state. Based on similar studies performed over SEIR dynamical systems some assumptions are taken into account Wu et al., (2020); Calafiore et al., (2020); Anastassopoulou et al., (2020). As of May 11, it is evident that how the early stage mathematical model parameters have changed drastically as detection rate was really low until middle of February, however the outbreak situation has improved comprehensively in several countries due to massive scale testing Qianying et al. (2020). We have considered the nominal values of the model parameters understanding the characteristics of the coronavirus infection, quantitatively estimated in the literature or published by health organizations Read et al., (2020); Li et al., (2020); Wu et al., (2020); Verity et al., (2020). In sequence, a rigorous process of model calibration which is known as Trust-region-reflective algorithm is applied to determine the best-fitted parameter values of the model. Our analysis was based on the publicly available data of the new confirmed daily cases reported for the future probable COVID-19 hotspots which are Russia, Brazil, India and Bangladesh from late January until May 09, 2020 Center for Systems Science and Engineering at Johns Hopkins University, (2020). Further, we have validated our model for the United Kingdom using same time frame. Based on the released data, we attempted to estimate the mean values of the crucial epidemiological parameters for COVID-19, such as the infectiousness factor for asymptotic carriers, isolation period, the estimation of the inflection point with probable date, recovery period for both symptomatic and asymptomatic individuals, case-fatality ratio and mortality rate in those potential hotspots. Perfect data-driven and curve-fitting methods for the prediction of any disease outbreak have always been question of interest in epidemiological research. Trust-region-reflect algorithm is one of the robust least-square data-fitting techniques that can promote a fine relation between the model driving mechanisms and model responses. This real-time data fitting approach could be efficient providing considerable insights on disease outbreak dynamics in different countries and designing worthwhile public health policies in curtailing the disease burden. Calibrating our model parameters by using the above algorithm, we have provided probable forecasts for the newly evolving COVID-19 hotspots.

This paper is organized as follows. In section 2, the mathematical model is described and the background of choosing baseline parameter values for the model is discussed. In section 3 and 4, the model has been analysed and model calibration technique has been discussed respectively. In section 5, model prediction accuracy has been illustrated by representing a graphical comparison between model responses and real-time data. In section 6, one of the robust global sensitivity analysis techniques is used to quantify the most influential mechanisms in our model. This paper ends with some qualitative and quantitative observations and discussions.

## 2. Formulation of the Mathematical Model

In this work, we use a compartmental differential equation model for the spread of COVID-19 in the world. The spread of the infection starts with the introduction of a small group of infected individuals to a large population. In this model we focus our study on eight components of the epidemic flow, i.e. {*S*(*t*), *E*(*t*), *I_D_*(*t*), *I_U_*(*t*), *Q*(*t*)*, H*(*t*), *R*(*t*)*, D*(*t*)} which represent the number of the susceptible individuals, exposed individuals (infected however not yet to be infectious, in an incubation period), symptomatic infectious individuals (confirmed with infectious capacity), asymptomatic infectious individuals (undetected but infectious), quarantined, hospitalized (under treatment), recovered cases (immuned) and death cases (or closed cases). Then the entire number of population in a certain region or country is *N* = *S* + *E* + *I_D_* + *I_U_* + *Q* + *H* + *R* + *D*. The following assumptions are considered in the formulation of the model:

- Emigration from the population and immigration into the population have not been taken into account in model formulation as there is negligible proportion of individuals move in and out of the population at a specific time frame.
- Births and natural deaths in the population are not considered.
- The susceptible population are exposed to a latent class.
- Hospitalized patients cannot spread disease while in isolation treatment after confirmed diagnosis.
- Recovered individuals do not return to susceptible class as they develop certain immunity against the disease. Hence they cannot become re-infected again and cannot infect susceptible either.

Figure 1 is the flow diagram of the proposed model, where susceptible individuals (S) get infected at a baseline infectious contact rate *β*, via contact with either symptomatic infectious individuals (*I_D_*) or at a rate *βλ* of asymptomatic infectious carriers and move to exposed (*E*) class. The relative infectiousness capacity of asymptomatic spreaders is λ (related to symptomatic infectious individuals). Susceptible individuals can be quarantined with contact tracing procedure and the rate of getting home-quarantined or self-quarantined of S is *q*, whereas κ is the rate of progression of symptoms of COVID-19 (hence 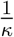 is the mean incubation period of COVID-19). Detected symptomatic infected individuals are generated at a progression rate *σ*_1_ and undetected asymptomatic infectious cases are generated at a rate *σ*_2_ from the exposed class. In addition, individuals who are exposed to virus can be quarantined with contact tracing procedure at a rate (1 − *σ*_1_ − *σ*_2_). *γ* is the rate of confirmation and isolation after symptom onset in detected infectious individuals and 1/*γ* is the time period when the infection can spread. The proportion of individuals from quarantined to symptomatic infectious, asymptomatic infectious and susceptible are *r*_2_*n*, *r*_1_*n* and (1− *r*_1_ − *r*_2_)*η* respectively. *δ_U_* is the disease-induced death rate for undetected asymptomatic patients and *δ_U_* is the death rate for hospitalized patients. *ϕ_D_*, *ϕ_U_* and *ϕ_H_* are recovery rates for the symptomatic patients, asymptomatic carriers and hospitalized patients respectively. In addition, 1/*ϕ_D_*, 1/*ϕ_U_* and 1/*ϕ_H_* represent the mean period of isolation for the detected symptomatic infectious individuals, undetected asymptomatic carriers and hospitalized patients. Based on these assumptions, the basic model structure for the transmission dynamics of COVID-19 is illustrated by the following deterministic framework of nonlinear differential equations (1).

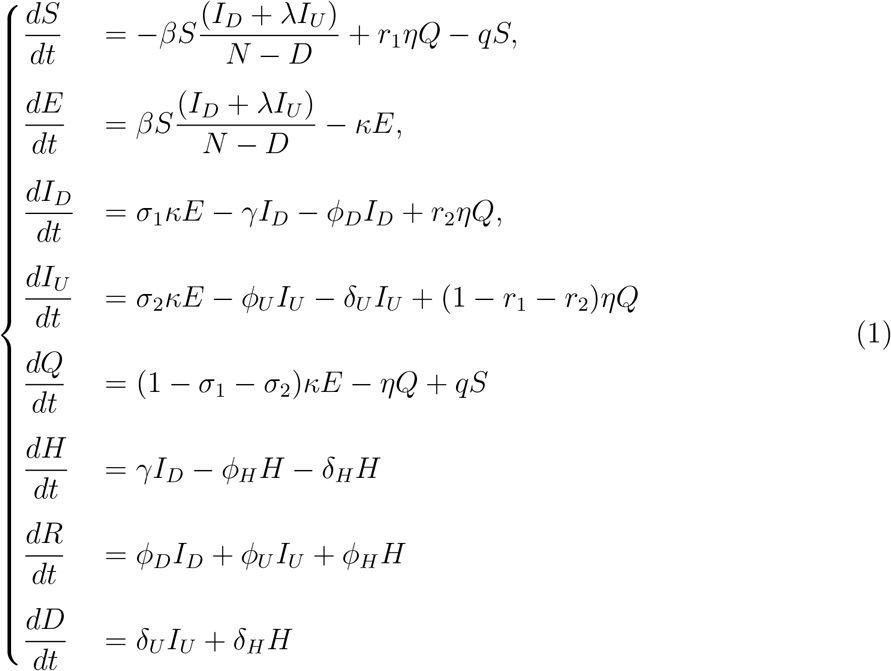

**Figure 1:**
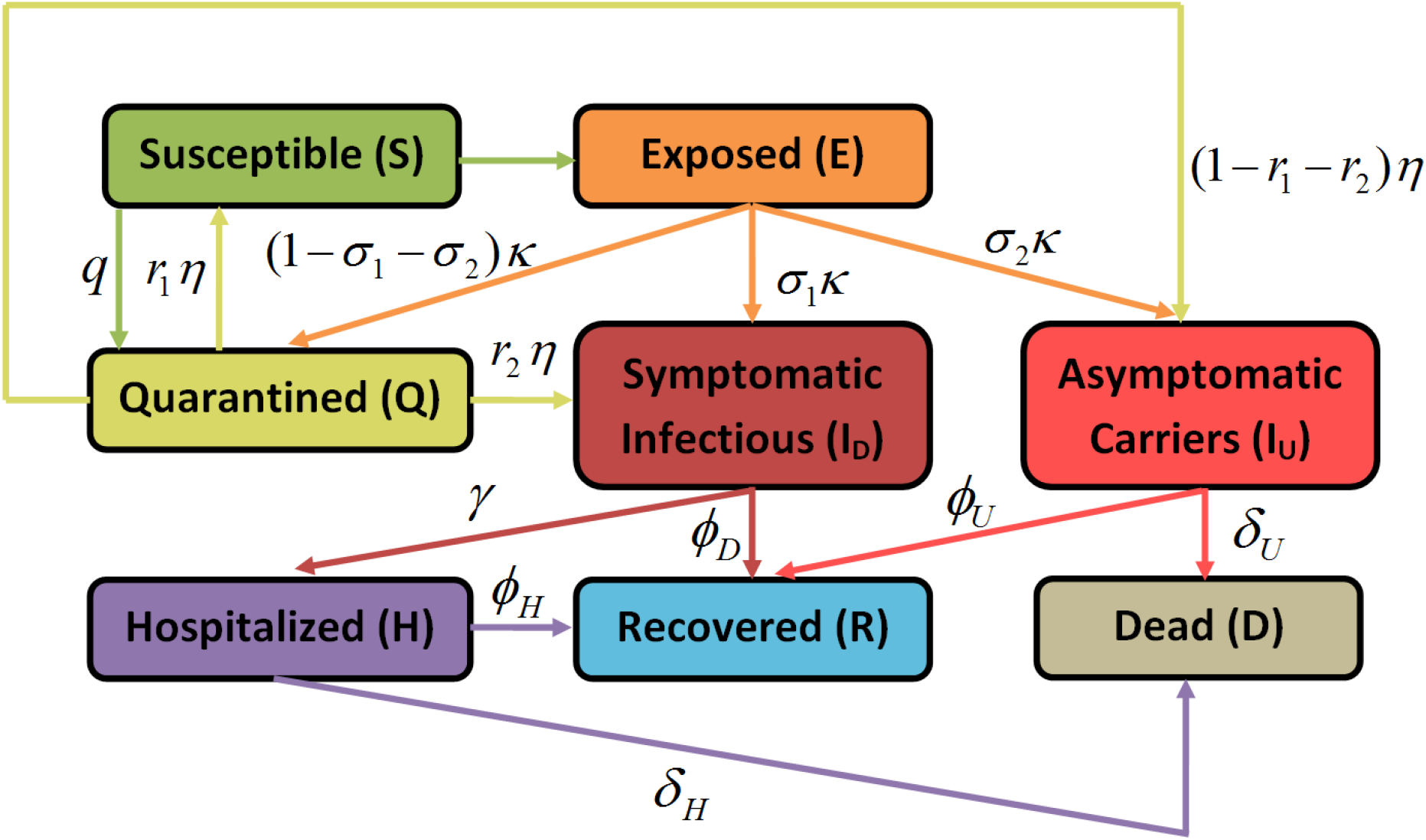
A schematic diagram that illustrates the proposed COVID-19 model

### 2.1. Baseline epidemiological parameters

In the study of Lauer et al., (2020), the average incubation period of COVID-19 was calculate to be 5.1 days and similar model-based studies also justified the above finding Li et al., (2020); Ferguson et al., (2020); Tang et al., (2020). Moreover, within 11.5 days (i.e. *κ* = 1/11.5 *day^−^*^1^) of infection, the individuals who were exposed to the virus started developing symptoms Lauer et al., (2020); Verity et al., (2020) and 20.0 days was the mean duration of viral shedding observed in COVID-19 survivors Zhou et al., (2020). The time duration from symptoms onset to recovery was estimated to be 24.7 days, whereas from onset of symptoms to death, the average mean duration was estimated to be 17Δ8 days Verity et al., (2020). In previous modeling studies, Shen et al. (2020); Read et al., (2020); Li et al., (2020), the novel coronavirus effective transmission rate, *β* ranges from around 0.25 − 1.5 *day^−^*^1^, which gradually follows a downward trend with time Tang et al., (2020). In sequence, we have considered this parameter as a time-varying parameter in our fits. Table 1 illustrates the list of baseline model parameters with brief description, probable ranges presented on clinical studies and calibration, and default base value selected for our study.

**Table 1:**
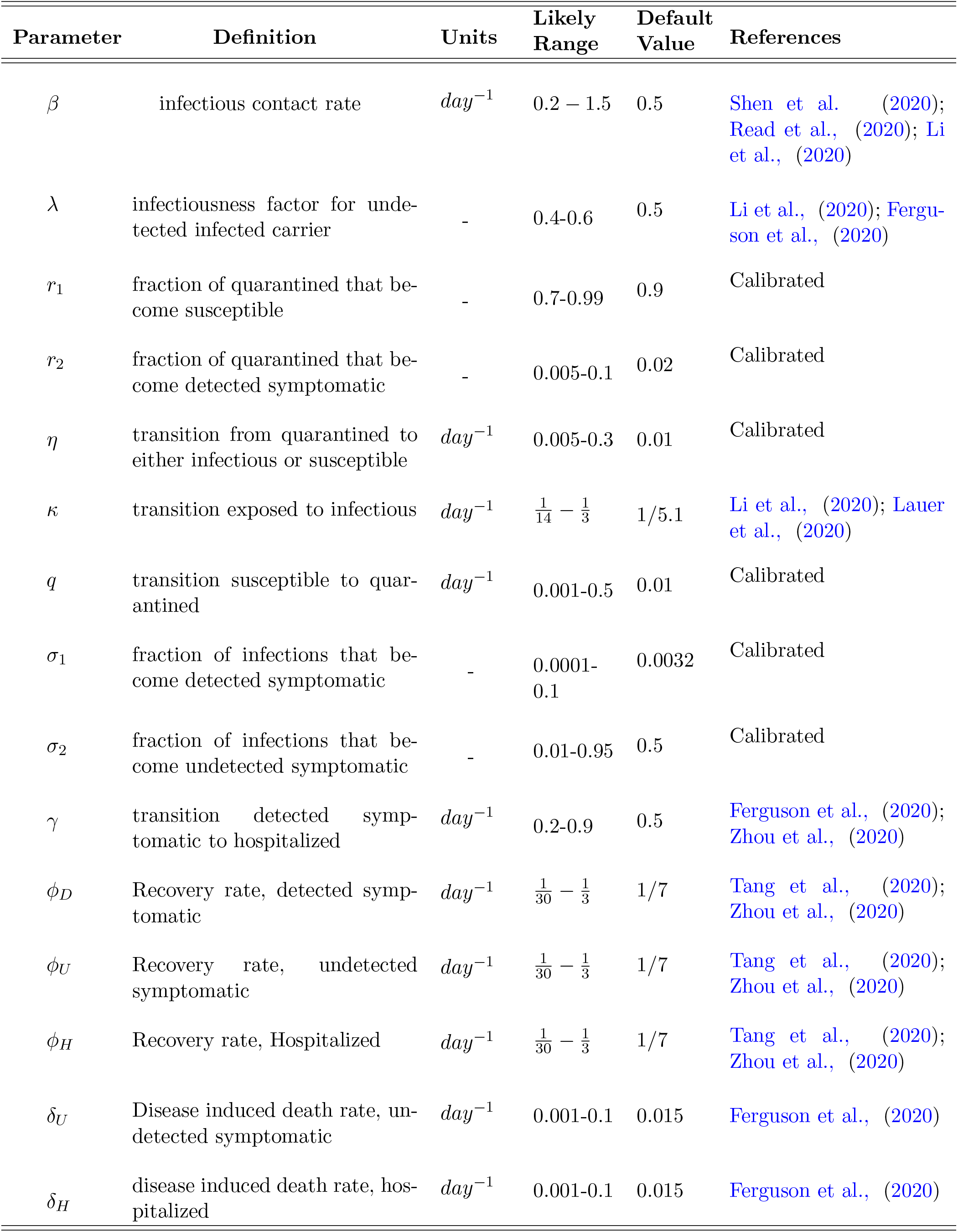
Model parameters with brief definition and probable ranges based on model calibration, relevant literature and clinical studies

### 2.2. Dataset

The recent epidemic data of five different countries which are Russia, Brazil, India, Bangladesh and the United Kingdom is accumulated from authoritative and genuine sources which are Center of Disease Control and Prevention (CDC) and the COVID Tracking Project (testing and hospitalizations). The data repository is handled by the Johns Hopkins University Center for Systems Science and Engineering (JHU CSSE) and supported by ESRI Living Atlas Team and the Johns Hopkins University Applied Physics Lab (JHU APL). The repository is publicly available and easy to compile Center for Systems Science and Engineering at Johns Hopkins University, (2020).

## 3. Analysis of the Model

### 3.1. Disease Free Equilibrium (DFE)

The disease-free equilibrium (DFE) of (1) can be obtained easily by setting *S* = *E* = *i_d_* = *I_U_* = *Q* = *H* = *R* = *D* = 0. Therefore the DFE of (1) is:

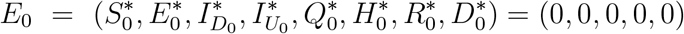

### 3.2. Basic Reproduction Number for Proposed Model

Using the next generation operator method Van den Driessche et al. (2002) the local stability of the DFE is investigated. According to the notation in Diekmann et al., (1990), the associated non-negative matrix, 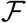 represents new infection terms, and the non-singular matrix, 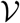 denotes remaining transfer termswhich can be described as follows:

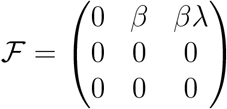

and

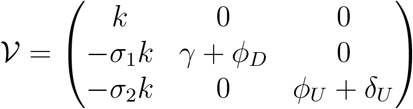

The associated basic reproduction number, denoted by 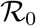 is then given by, 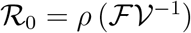, where *ρ* is the spectral radius of 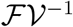. It follows that,

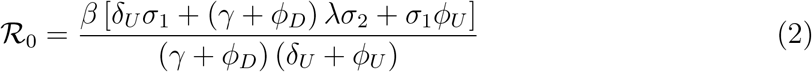

Thus, by Theorem 2 of LaSalle (1976), the following result is established.

#### Lemma 1.

*The DFE, E_0_ of the system* (1)*, is locally-asymptotically stable* (*LAS*) *if* 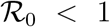, *and unstable if* 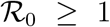

The threshold quantity, 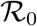, estimates the mean number of secondary cases generated by a single infected individual in an entirely susceptible human population Hethcote (2000). The above result implies that a small influx of infected individuals would not generate large outbreaks if 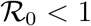, and the disease will persist (be endemic) in the population if 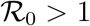.

## 4. Model Calibration

The proposed epidemic model (1) is a continuous-time non-linear system of differential equations together with a suitable set of initial conditions. In this study, Trust-region-reflective (TRR) algorithm has been used to determine the best-fitted parameters for our proposed model. We have used lsqcurvefit function in MATLAB to calibrate our model. The optimization process can be expressed as follows:

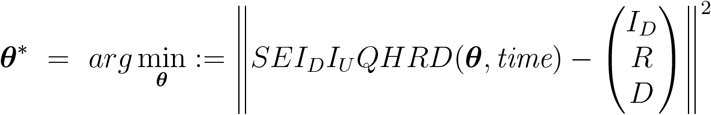

where ***θ***\* is the set of parameters of dynamically calibrated model, {*I_D_*, *R*, *D*} is a set of the detected symptomatic infectious individuals, recovered and disease induced death cases from the real-time data and ***θ*** = {*β*, *λ*, *r*_1_, *r*_2_, *η*, *κ, σ*_1_*, σ*_2_*, γ, q, ϕ_D_, ϕ_U_, ϕ_H_, δ_U_, δ_H_ }* is the initial set of parameters of the proposed model and *SEI_D_I_U_QHRD*(.) represents our proposed model. The “TRR algorithm” also necessitates an initial guess for each parameter which is described as “TRR input” in the following section.

## 5. Numerical Experiments and Forecasting

In this section, we have used dynamically calibrated compartment model for real-time analysis and real-time prediction of COVID-19 outbreak for five different countries which are Russia, Brazil, India, Bangladesh and the United Kingdom. Figure 2, 4, 6, 8, 10 illustrates the best real-time data fitting results using the baseline parameters from Table 1 as initial inputs for our proposed model. The ranges of the parameters and have been set compatible with the clinical studies and illustrated 2, 3, 4, 5, 6 along with the parameters resulted from the calibration (“TRR output”). To ensure a huge number of susceptible individuals at the initial stage of an outbreak, 0.9 *N* has been set as a lower bound for *S*.

**Figure 2:**
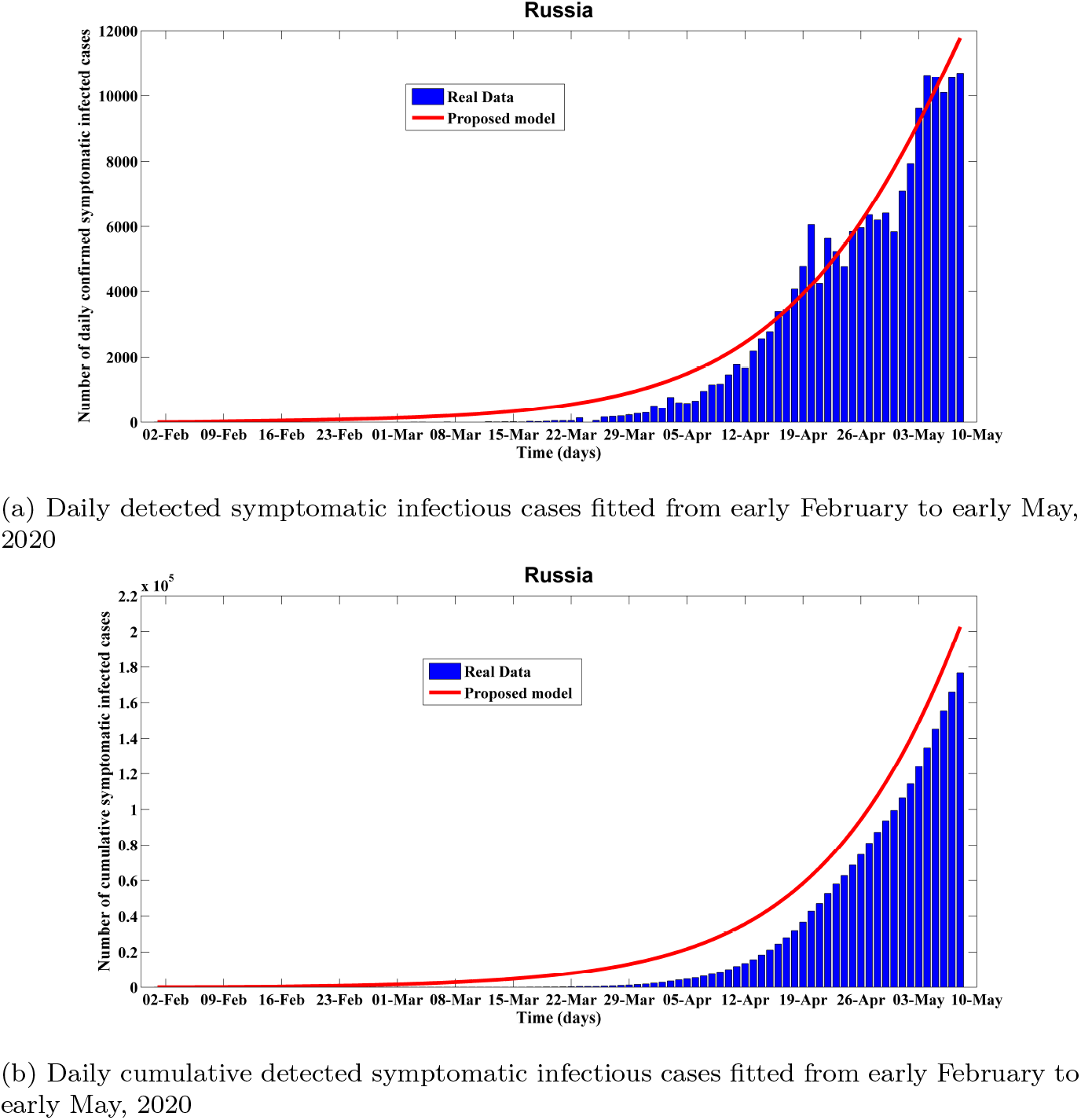
Fitting performance of calibrated *sei_d_i_u_qhrd* model for Russia from February 1 to May 08, 2020

### 5.1. Analysis and prediction for Russia

With much of Europe now easing itself out of confinement, Russia could become the continent’s new Covid-19 hotspot according to our analysis. The model fitting and projection results for Russia from early February to late August are shown in Figure 2 and 3. We collected real data from February 01 to May 08, 2020 to calibrate the model parameters.

**Figure 3:**
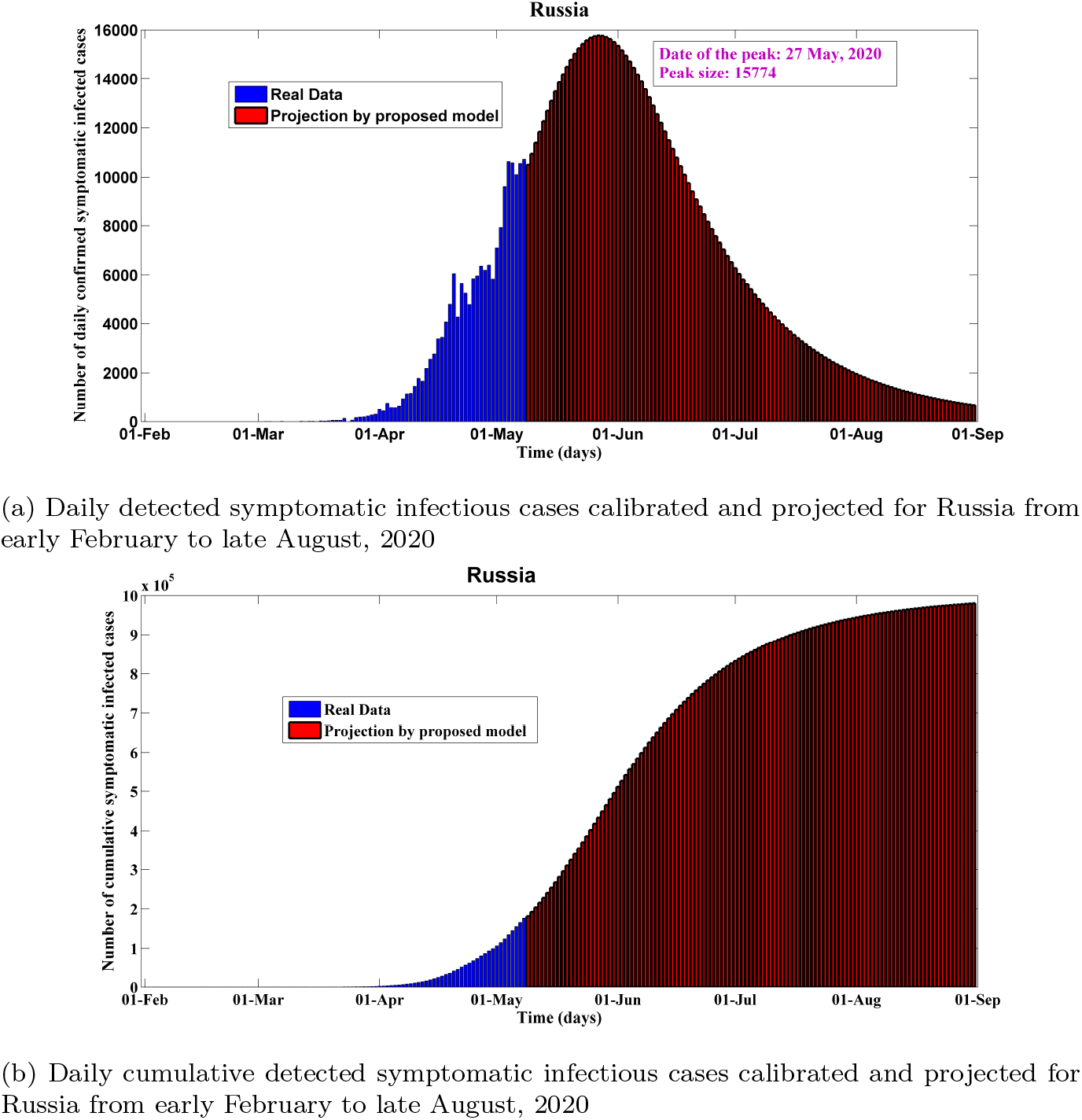
Predictions of the proposed *sei_d_i_u_qhrd* model for Russia from early March to late August, 2020

**Figure 4:**
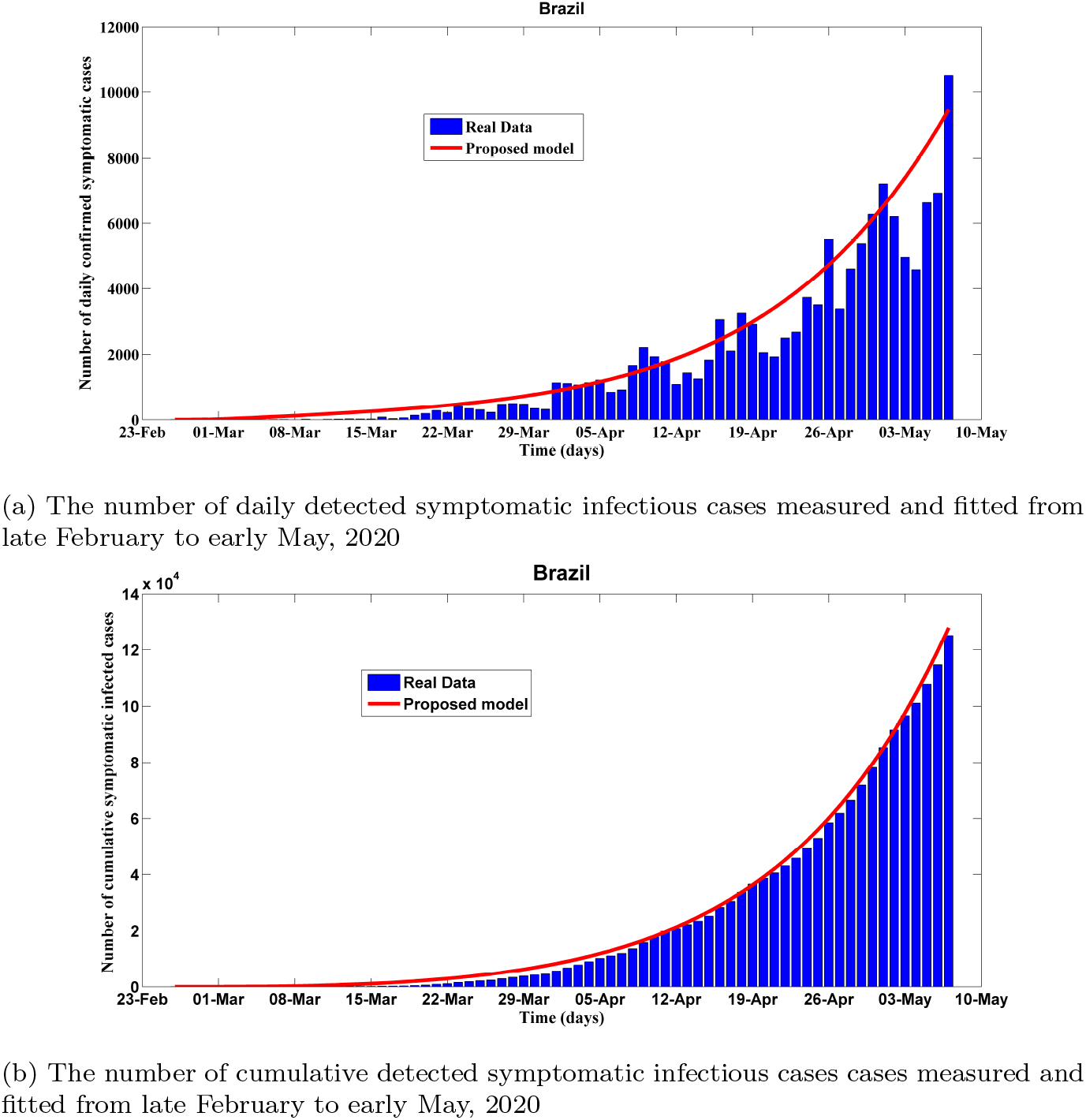
Fitting performance of calibrated *sei_d_i_u_qhrd* model for Brazil from 26 February to 8 May, 2020

As we can see the results from the proposed model match the real data very well. Based on the proposed model, we want to project that from Figure 3, the number of daily detected symptomatic infectious cases in Russia will reach the peak at May 27 with about 15.774K cases. As time progresses, our estimated daily projected mean error drops to 10% for the cumulative cases and daily new cases which represents the robustness of the model forecasting. The basic reproduction number is 4.234 as of May 08, which lies in prior established findings 2 − 7 for COVID-19 Liu et al., (2020). The number of cumulative infected cases is projected to reach 970K around August 30, and the estimated total death cases will reach to about 35K in the end. To date, Russia’s official death toll of 1,827 is relatively low because not all deaths of people who have contracted the virus are being counted as Covid-19 deaths. Table 2 illustrates the key features used to calibrate this scenario which are compatible with the previous clinical studies and relevant literature.

**Table 2:**
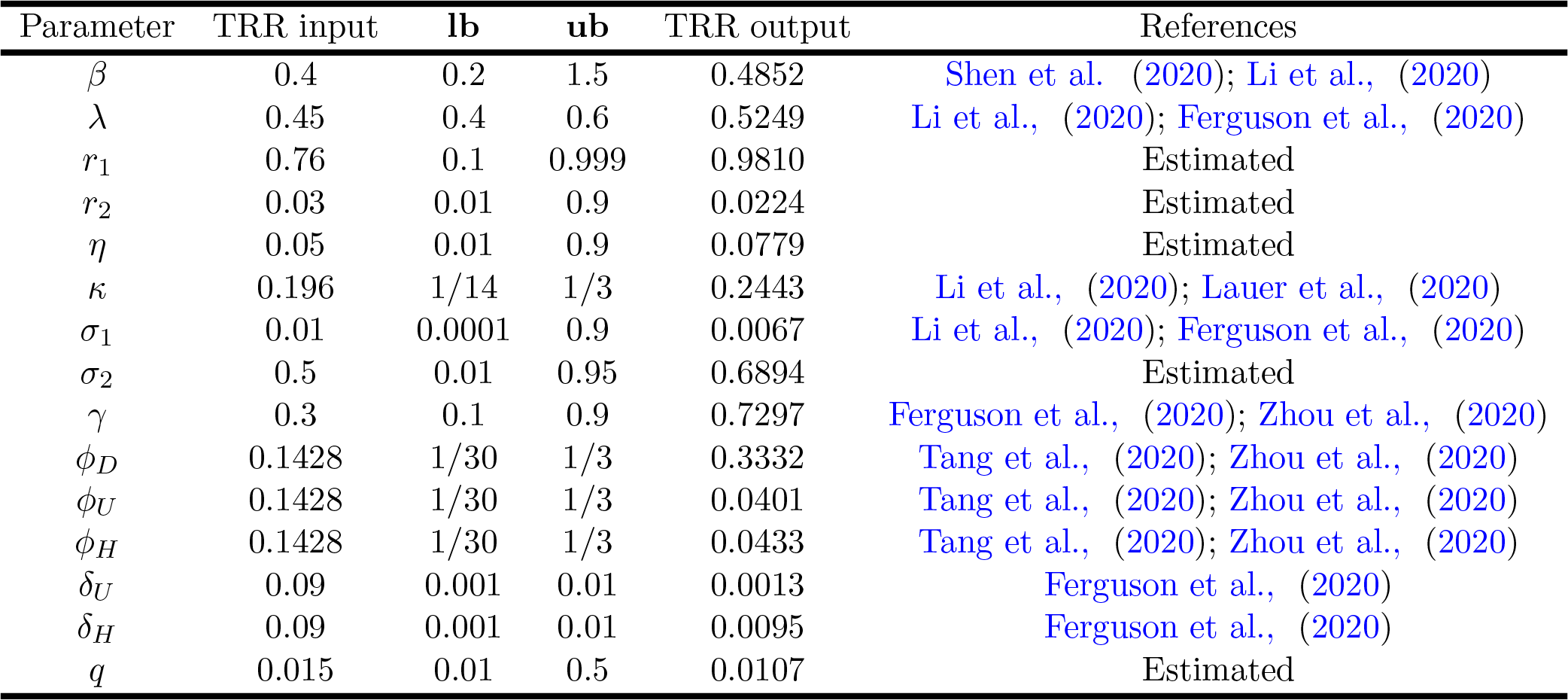
Necessary parameters for trust-region-reflective algorithm and for the Figure 3 calibrated response

| Parameter | TRR input | lb | ub | TRR output | References |
| --- | --- | --- | --- | --- | --- |
| $\beta$ | 0.4 | 0.2 | 1.5 | 0.4852 | Shen et al., (2020); Li et al., (2020) |
| $\lambda$ | 0.45 | 0.4 | 0.6 | 0.5249 | Li et al., (2020); Ferguson et al., (2020) |
| $r_1$ | 0.76 | 0.1 | 0.999 | 0.9810 | Estimated |
| $r_2$ | 0.03 | 0.01 | 0.9 | 0.0224 | Estimated |
| $\eta$ | 0.05 | 0.01 | 0.9 | 0.0779 | Estimated |
| $\kappa$ | 0.196 | 1/14 | 1/3 | 0.2443 | Li et al., (2020); Lauer et al., (2020) |
| $\sigma_1$ | 0.01 | 0.0001 | 0.9 | 0.0067 | Li et al., (2020); Ferguson et al., (2020) |
| $\sigma_2$ | 0.5 | 0.01 | 0.95 | 0.6894 | Estimated |
| $\gamma$ | 0.3 | 0.1 | 0.9 | 0.7297 | Ferguson et al., (2020); Zhou et al., (2020) |
| $\phi_D$ | 0.1428 | 1/30 | 1/3 | 0.3332 | Tang et al., (2020); Zhou et al., (2020) |
| $\phi_U$ | 0.1428 | 1/30 | 1/3 | 0.0401 | Tang et al., (2020); Zhou et al., (2020) |
| $\phi_H$ | 0.1428 | 1/30 | 1/3 | 0.0433 | Tang et al., (2020); Zhou et al., (2020) |
| $\delta_U$ | 0.09 | 0.001 | 0.01 | 0.0013 | Ferguson et al., (2020) |
| $\delta_H$ | 0.09 | 0.001 | 0.01 | 0.0095 | Ferguson et al., (2020) |
| $q$ | 0.015 | 0.01 | 0.5 | 0.0107 | Estimated |

### 5.2. Analysis and prediction for Brazil

The coronavirus disease 2019 (COVID-19) pandemic headed toward Latin America later than other continents. On Feb 25, 2020, the first infected case was documented. But now, Brazil has surpassed the records in Latin America in terms of deaths and new infected cases (155939 cases and 10627 deaths as of May 9). This is probably an underestimated scenario in comparison to real severity in Brazil. Our analysis projects that Brazil is developing as one of the world’s next coronavirus hotspots. The model fitting and projection results for Brazil from late February to late August are shown in Figure 4 and 5. We took real data from February 25 to May 08 to calibrate the model parameters. As we can see the results from the proposed model fit the real data very well. Based on the proposed model, we project that from Figure 5, the number of daily detected symptomatic infectious cases in Brazil seem reaching the peak around June 11 with about 26.449K cases. The basic reproduction number is estimated about 5.3467 as of May 11, which is in between the observed basic reproduction number for COVID-19, estimated about 2-7 for COVID-19 Liu et al., (2020). This crucial epidemiological parameter could blow up because of scant diagnostics and impermanent non-pharmaceutical interventions. The case-fatality rate is hovering around 9.3% as of May 11, which is necessary to assess how much community transmission has occurred and its burden. According to our projection, this ratio could be doubled within two months. The number of cumulative infected cases is projected to reach 1800K around August 30 if current trend is held, and the estimated total death cases will reach to about 108K in the end. Table 3 illustrates the key features used to calibrate this scenario which are compatible with the previous clinical studies and relevant literature.

**Figure 5:**
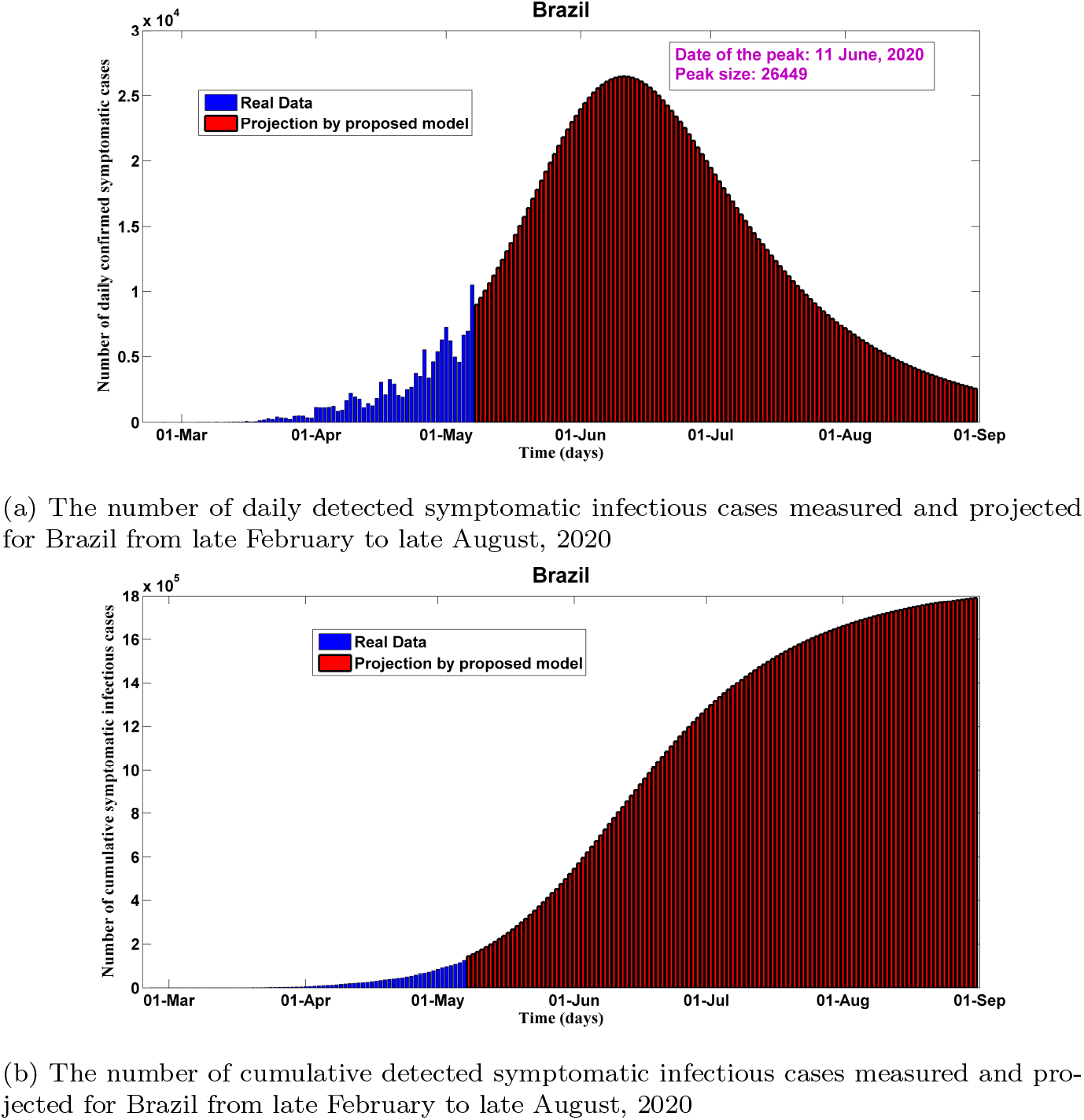
Predictions of the proposed *SEI_D_I_U_QHRD* model for Brazil from late February to late August, 2020

**Figure 6:**
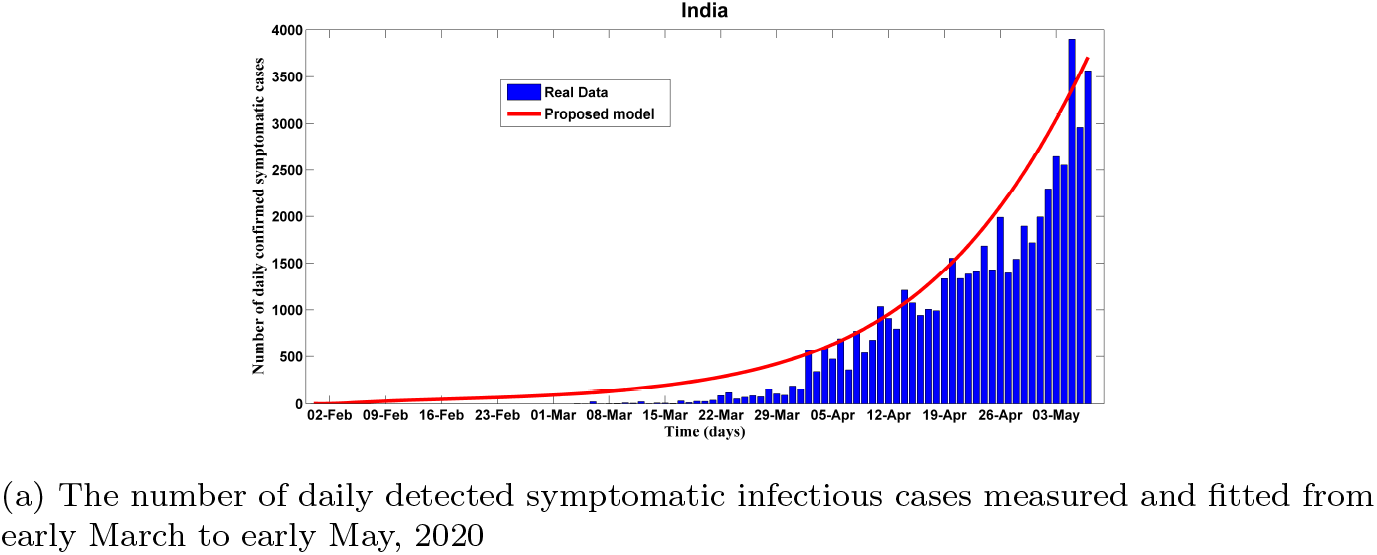
Fitting performance of calibrated *SEI_D_I_U_QHRD* model for India from 8 March to early May, 2020

**Table 3:**
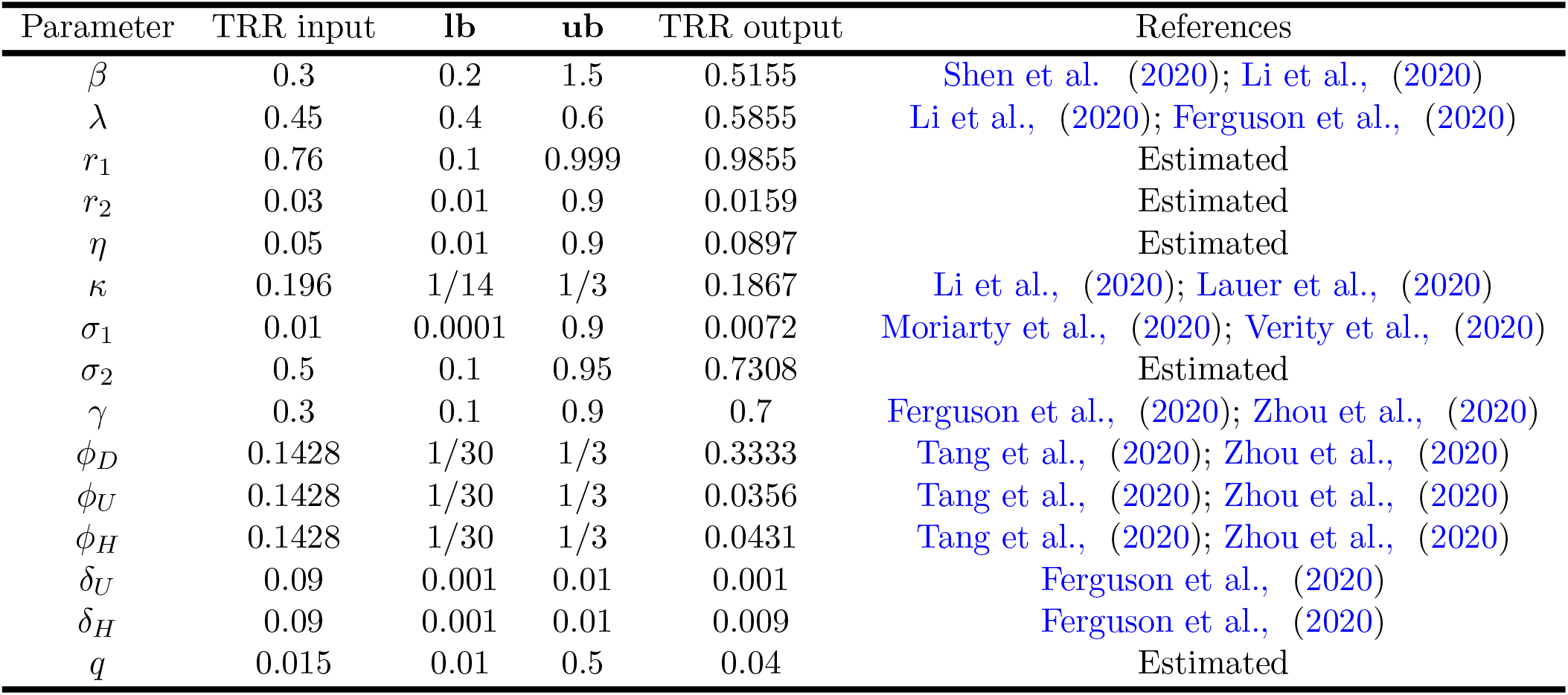
Necessary parameters for trust-region-reflective algorithm and for the Figure 5 calibrated response

### 5.3. Analysis and prediction for India

Some people might think about the possibilities of the presence of a less virulent strain of the virus in India, along with the possibility that its hot weather could diminish the contagion. Nevertheless, our mathematical analysis suggests that India could become the new Covid-19 hotspot in South Asia. According to our projection, India could continue seeing spikes in the number of cases as it time progresses despite of following lockdown and other disease mitigation measures. The first case of the COVID-19 pandemic in India was reported on 30 January 2020, originating from China. But now, India has the most cases and deaths in South Asia (62,808 cases and 2,101 deaths as of May 9), and these are probably substantial underestimates. Figure 6 and 7 illustrate COVID-19 disease modeling and prediction for India from late January to late August. We took real-time data from January 30 to May 8 to calibrate the model parameters. As we can see, our proposed model match well for the historical real data. Based on our prediction from Figure 7, the number of daily detected symptomatic infectious cases in India seem reaching the peak around June 15 with about 9.504K cases. The basic reproduction number is 5.218 as of May 11, which lies between the studied observations Liu et al., (2020). This estimation may be considered as an overestimation of this critical parameter. Notwithstanding, this is owing to the infectiousness factor of the asymptomatic spreaders. Minimally or reasonably symptomatic infectious and the asymptomatic spreaders should be quarantined to avoid the transmission of the virus, with the critically symptomatic patients isolated in proper health care settings. By carrying out massive scale contract tracing, risky individuals could be identified by means of their exposure. The number of cumulative infected cases is projected to reach 730K around August 30 if current pattern is held, and the estimated total death cases will reach to about 43.8K in the end. Table 4 illustrates the key features used to calibrate this scenario which are compatible with the previous clinical studies and relevant literature. Interestingly, we have found in our analysis that India’s case-fatality rate is at 3.5% and the country’s recovery rate is at 33% which commensurate with real reported statistics precisely.

**Figure 7:**
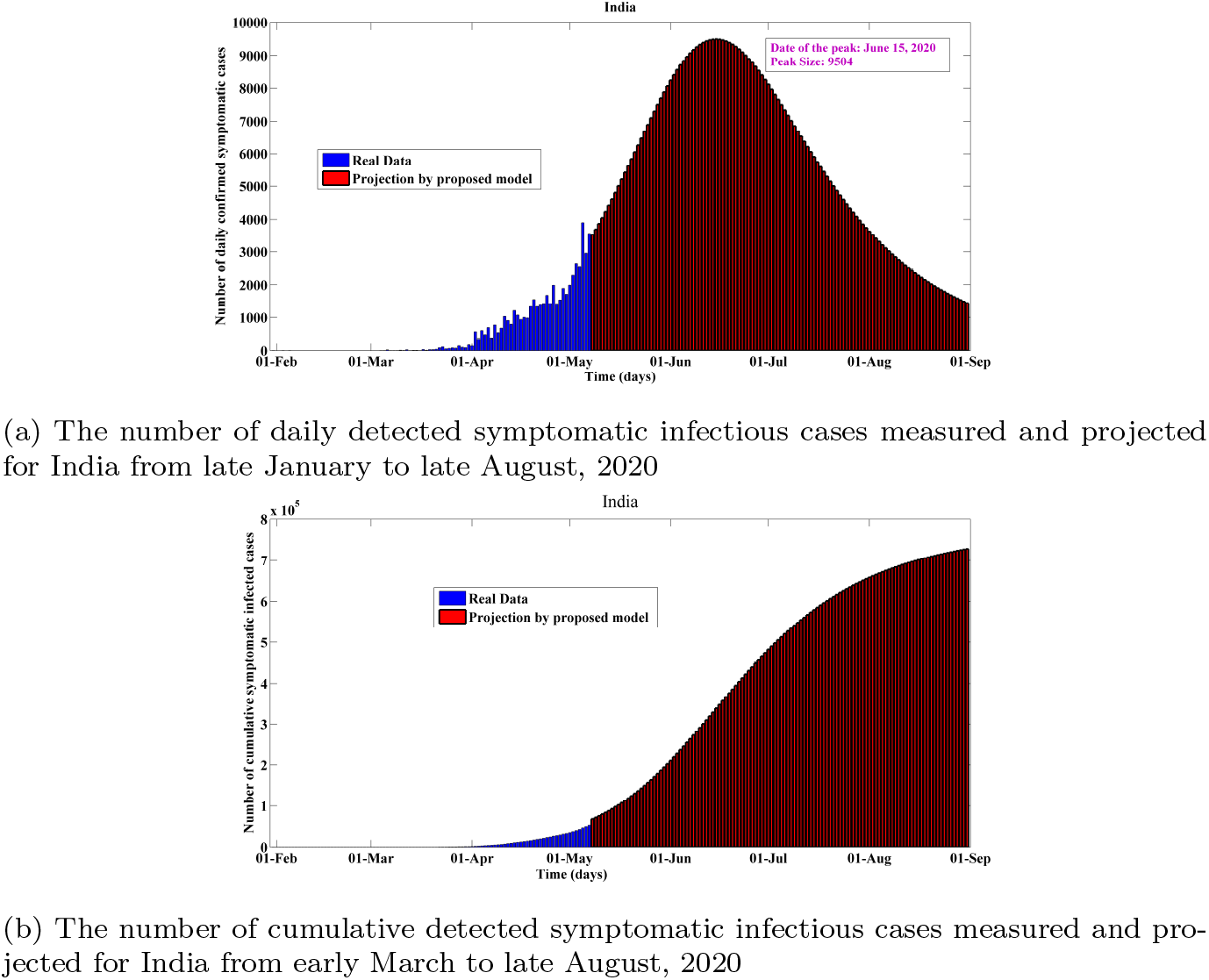
Predictions of the proposed *sei_d_i_u_qhrd* model for India from late January to late August, 2020

**Figure 8:**
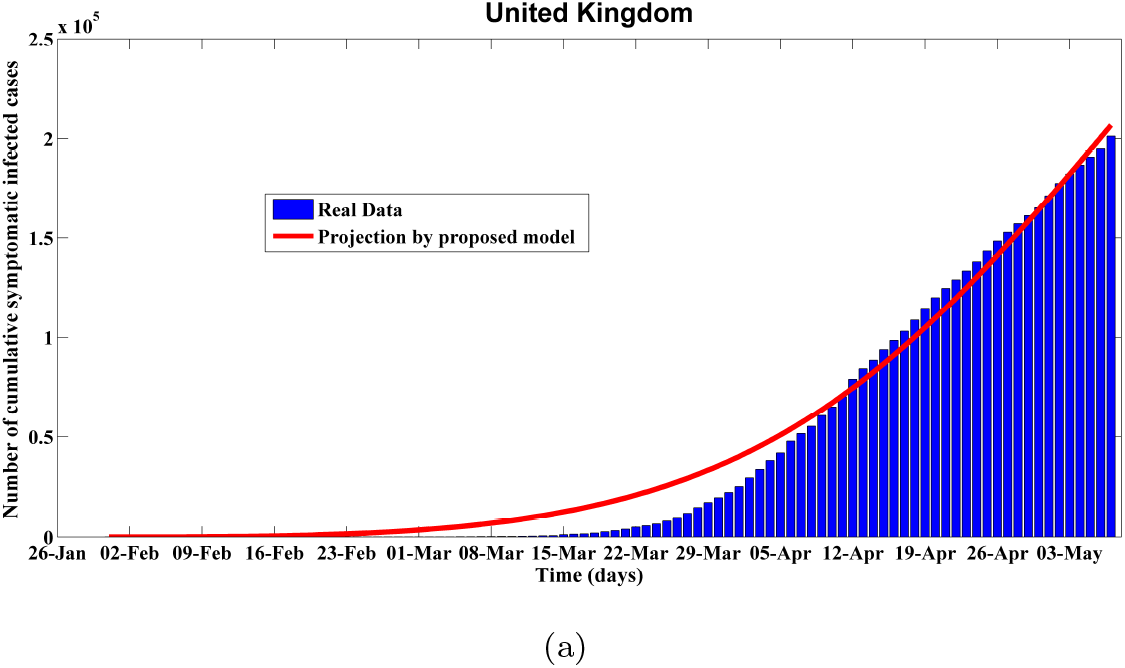
Fitting performance of calibrated *SEI_D_I_U_QHRD* model for the United Kingdom from 31 January to early May, 2020

**Table 4:** Necessary parameters for trust-region-reflective algorithm and for the Figure 7 calibrated response

| Parameter | TRR input | lb | ub | TRR output | References |
| --- | --- | --- | --- | --- | --- |
| $\beta$ | 0.3 | 0.2 | 1.5 | 0.3995 | Shen et al., (2020); Li et al., (2020) |
| $\lambda$ | 0.4 | 0.4 | 0.6 | 0.5995 | Li et al., (2020); Ferguson et al., (2020) |
| $r_1$ | 0.76 | 0.1 | 0.999 | 0.9911 | Estimated |
| $r_2$ | 0.03 | 0.01 | 0.9 | 0.01 | Estimated |
| $\eta$ | 0.05 | 0.01 | 0.9 | 0.0997 | Estimated |
| $\kappa$ | 0.196 | 1/14 | 1/3 | 0.3296 | Li et al., (2020); Lauer et al., (2020) |
| $\sigma_1$ | 0.01 | 0.0001 | 0.1 | 0.00046 | Moriarty et al., (2020); Verity et al., (2020) |
| $\sigma_2$ | 0.5 | 0.1 | 0.95 | 0.7495 | Estimated |
| $\gamma$ | 0.3 | 0.1 | 0.9 | 0.8 | Ferguson et al., (2020); Zhou et al., (2020) |
| $\phi_D$ | 0.1428 | 1/30 | 1/3 | 0.3333 | Tang et al., (2020); Zhou et al., (2020) |
| $\phi_U$ | 0.1428 | 1/30 | 1/3 | 0.0334 | Tang et al., (2020); Zhou et al., (2020) |
| $\phi_H$ | 0.1428 | 1/30 | 1/3 | 0.3335 | Tang et al., (2020); Zhou et al., (2020) |
| $\delta_U$ | 0.09 | 0.001 | 0.01 | 0.001 | Ferguson et al., (2020) |
| $\delta_H$ | 0.09 | 0.001 | 0.01 | 0.01 | Ferguson et al., (2020) |
| $q$ | 0.015 | 0.01 | 0.5 | 0.0507 | Estimated |

### 5.4. Analysis and prediction for the United Kingdom

Our analysis indicates that the United Kingdom could face risk of a second wave of coronavirus infections as the United Kingsom gradually eases a nationwide lockdown. According to Worldometer, (2020), by 11 May there had been 223,060 confirmed cases and 32,065 deaths overall a rate of 465 deaths per million population. The outbreak in London has the highest number and highest rate of infections, while England and Wales are the UK countries with the highest recorded death rate per capita. Recently, The prime minister of the UK has expressed his plan to unveil a coronavirus warning system as an intervention strategy, while he was planning to ease the lockdown gradually in the UK. For the UK, the modeling and projection results from January 31 to August 31 are shown in Figure 8 and 9. As we can see from Figure 9, the number of daily detected symptomatic infectious cases reached the peak around April 10. Since then the curve has been maintaining a plateau which is a really unusual scenario. This phenomenon has closely been captured by our proposed model. This case study was really important for the validation of our model. In fact, this guarantees the fact that this model is capable of providing more precise and vigorous short-term predictions of COVID-19 dynamics. According to our calculation, the basic reproduction number is around 4.649 as of May 09, which lies in the prior studies of COVID-19 Anastassopoulou et al., (2020); Liu et al., (2020). The number cumulative infected cases is projected to reach 618K around August 30, and the estimated total death cases will reach to about 63.48K in the above mentioned period. Importantly, as of May 11, we have found in our analysis that the UK’s case-fatality rate is at 17.2%, which is the worst among our five studied cases. This could exacerbate as time progresses in the absence of proven effective therapy or a vaccine. In addition, Our study suggests that relaxing social distancing too soon could result in thousands of additional death in the UK. Table 5 illustrates the key features used to calibrate this scenario which are compatible with the previous clinical studies and relevant literature.

**Figure 9:**
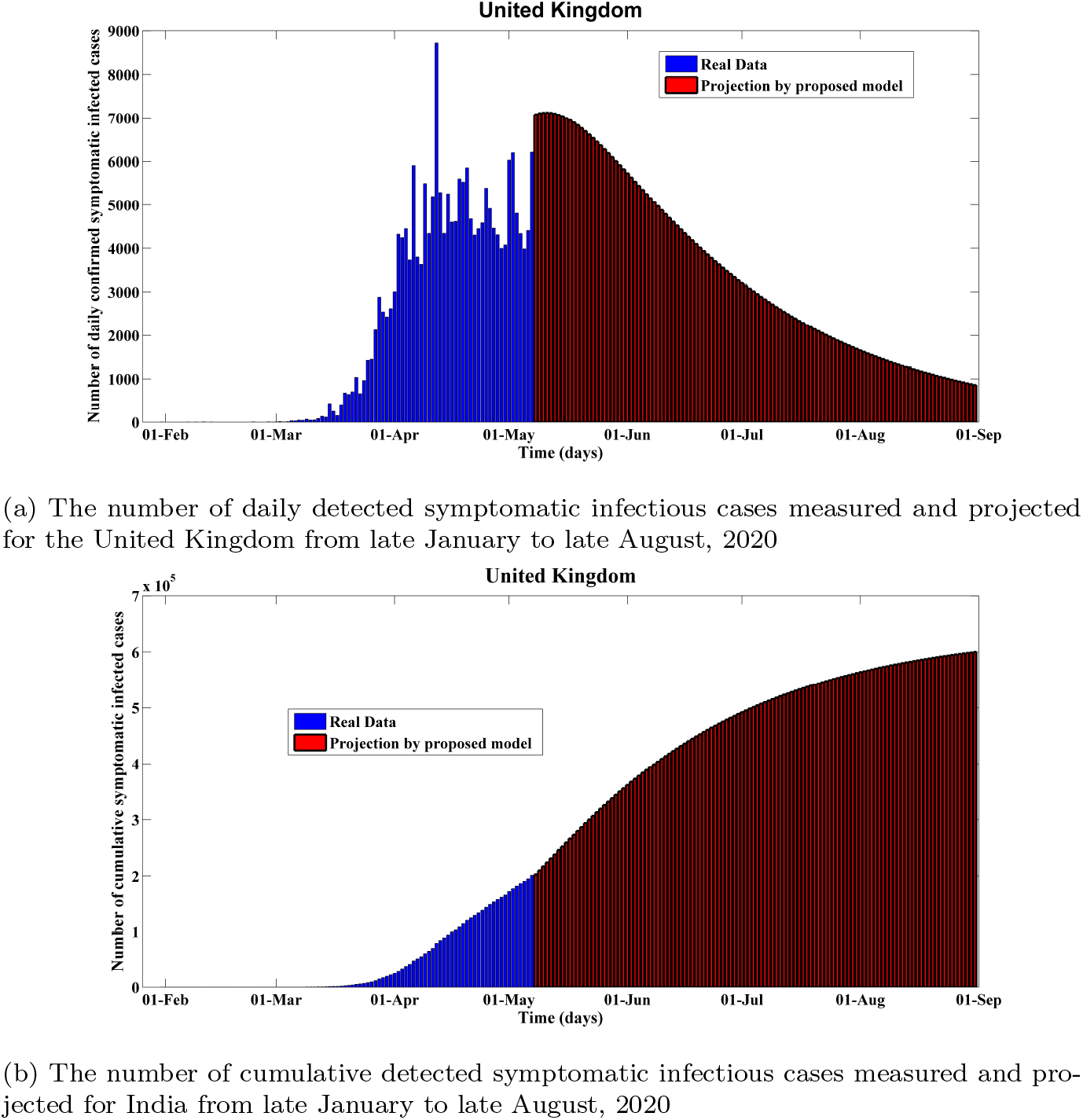
Predictions of the proposed *sei_d_i_u_qhrd* model for the United Kingdom from late January to late August, 2020

**Table 5:**
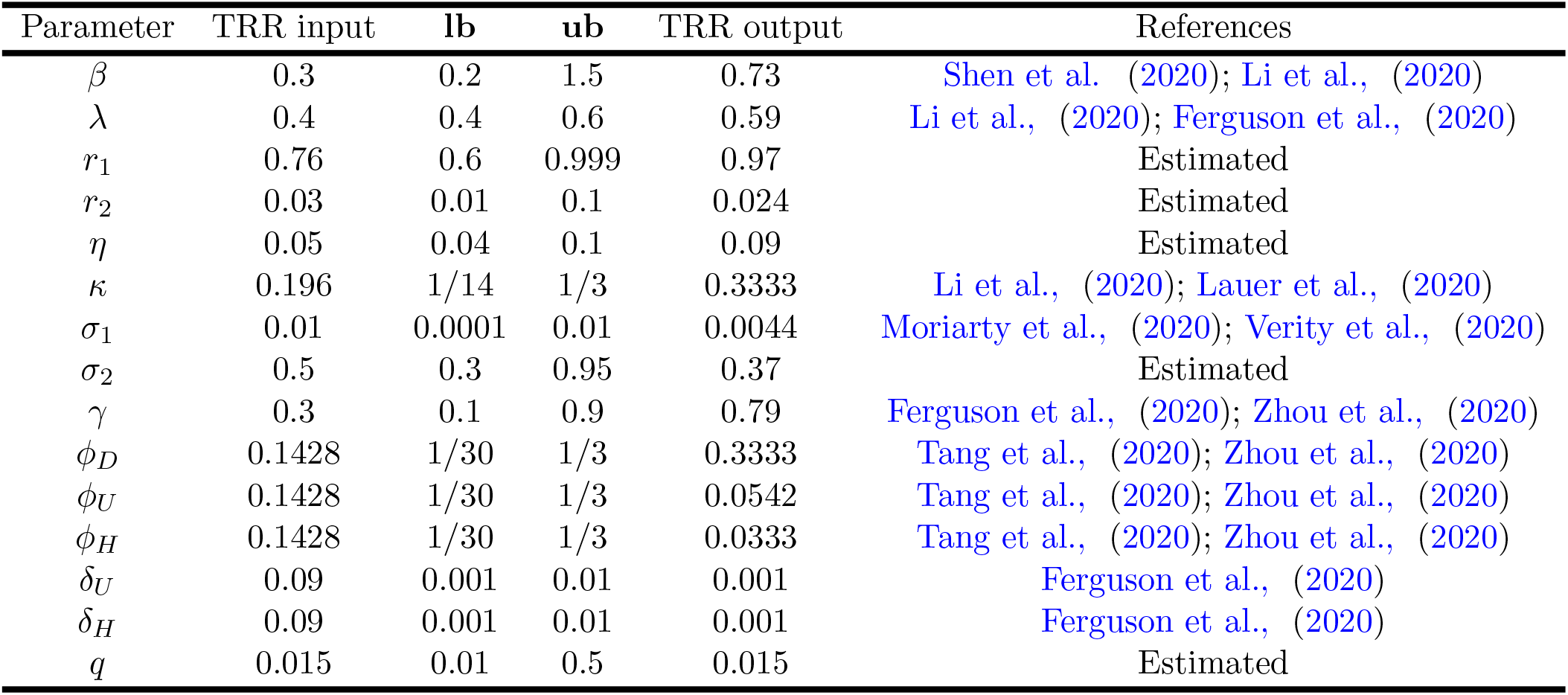
Necessary parameters for trust-region-reflective algorithm and for the Figure 9 calibrated response

| Parameter | TRR input | lb | ub | TRR output | References |
| --- | --- | --- | --- | --- | --- |
| $\beta$ | 0.3 | 0.2 | 1.5 | 0.73 | Shen et al. (2020); Li et al., (2020) |
| $\lambda$ | 0.4 | 0.4 | 0.6 | 0.59 | Li et al., (2020); Ferguson et al., (2020) |
| $r_1$ | 0.76 | 0.6 | 0.999 | 0.97 | Estimated |
| $r_2$ | 0.03 | 0.01 | 0.1 | 0.024 | Estimated |
| $\eta$ | 0.05 | 0.04 | 0.1 | 0.09 | Estimated |
| $\kappa$ | 0.196 | 1/14 | 1/3 | 0.3333 | Li et al., (2020); Lauer et al., (2020) |
| $\sigma_1$ | 0.01 | 0.0001 | 0.01 | 0.0044 | Moriarty et al., (2020); Verity et al., (2020) |
| $\sigma_2$ | 0.5 | 0.3 | 0.95 | 0.37 | Estimated |
| $\gamma$ | 0.3 | 0.1 | 0.9 | 0.79 | Ferguson et al., (2020); Zhou et al., (2020) |
| $\phi_D$ | 0.1428 | 1/30 | 1/3 | 0.3333 | Tang et al., (2020); Zhou et al., (2020) |
| $\phi_U$ | 0.1428 | 1/30 | 1/3 | 0.0542 | Tang et al., (2020); Zhou et al., (2020) |
| $\phi_H$ | 0.1428 | 1/30 | 1/3 | 0.0333 | Tang et al., (2020); Zhou et al., (2020) |
| $\delta_U$ | 0.09 | 0.001 | 0.01 | 0.001 | Ferguson et al., (2020) |
| $\delta_H$ | 0.09 | 0.001 | 0.01 | 0.001 | Ferguson et al., (2020) |
| $q$ | 0.015 | 0.01 | 0.5 | 0.015 | Estimated |

**Table 6:**
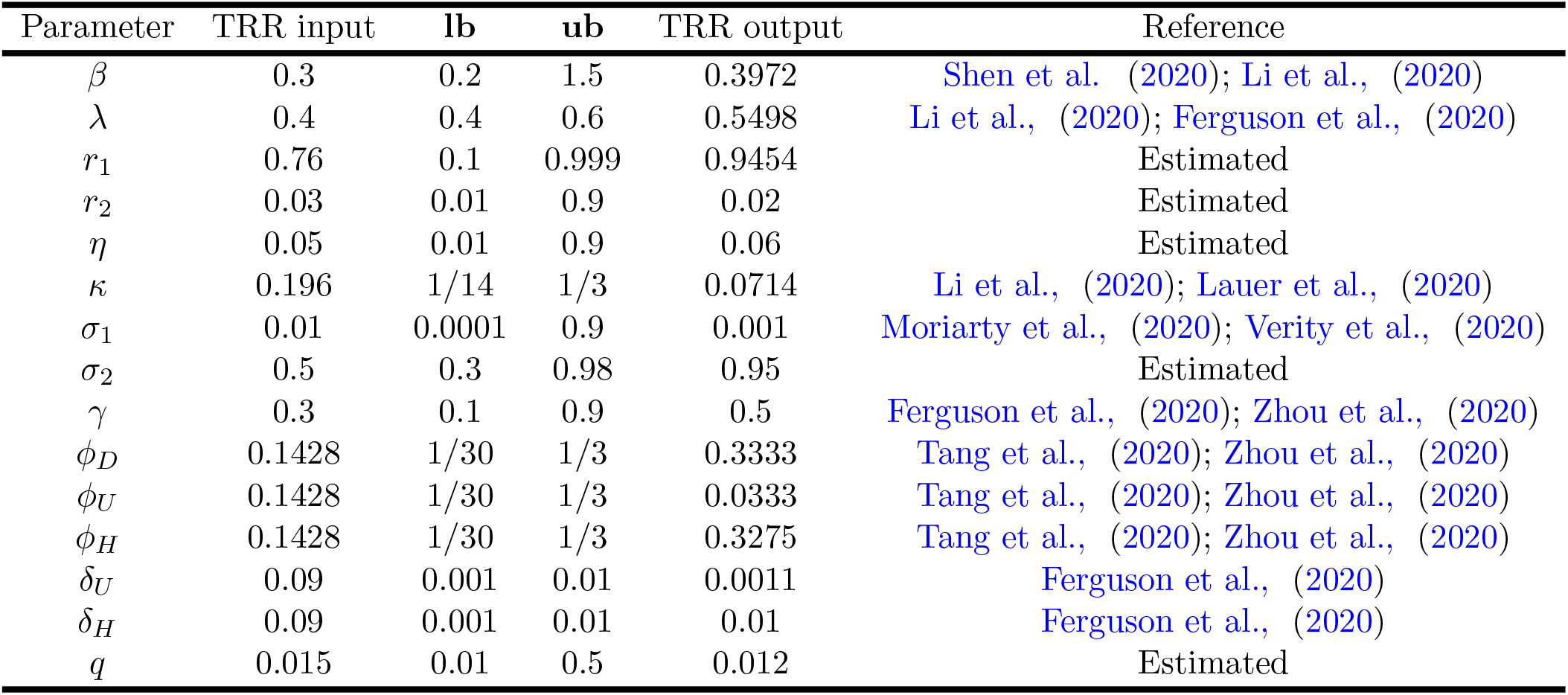
Necessary parameters for trust-region-reflective algorithm and for the Figure 11 calibrated response

| Parameter | TRR input | lb | ub | TRR output | Reference |
| --- | --- | --- | --- | --- | --- |
| $\beta$ | 0.3 | 0.2 | 1.5 | 0.3972 | Shen et al. (2020); Li et al., (2020) |
| $\lambda$ | 0.4 | 0.4 | 0.6 | 0.5498 | Li et al., (2020); Ferguson et al., (2020) |
| $r_1$ | 0.76 | 0.1 | 0.999 | 0.9454 | Estimated |
| $r_2$ | 0.03 | 0.01 | 0.9 | 0.02 | Estimated |
| $\eta$ | 0.05 | 0.01 | 0.9 | 0.06 | Estimated |
| $\kappa$ | 0.196 | 1/14 | 1/3 | 0.0714 | Li et al., (2020); Lauer et al., (2020) |
| $\sigma_1$ | 0.01 | 0.0001 | 0.9 | 0.001 | Moriarty et al., (2020); Verity et al., (2020) |
| $\sigma_2$ | 0.5 | 0.3 | 0.98 | 0.95 | Estimated |
| $\gamma$ | 0.3 | 0.1 | 0.9 | 0.5 | Ferguson et al., (2020); Zhou et al., (2020) |
| $\phi_D$ | 0.1428 | 1/30 | 1/3 | 0.3333 | Tang et al., (2020); Zhou et al., (2020) |
| $\phi_U$ | 0.1428 | 1/30 | 1/3 | 0.0333 | Tang et al., (2020); Zhou et al., (2020) |
| $\phi_H$ | 0.1428 | 1/30 | 1/3 | 0.3275 | Tang et al., (2020); Zhou et al., (2020) |
| $\delta_U$ | 0.09 | 0.001 | 0.01 | 0.0011 | Ferguson et al., (2020) |
| $\delta_H$ | 0.09 | 0.001 | 0.01 | 0.01 | Ferguson et al., (2020) |
| $q$ | 0.015 | 0.01 | 0.5 | 0.012 | Estimated |

### 5.5. Analysis and prediction for Bangladesh

For Bangladesh, the calibration and projection results from early March to late August of are shown in Figure 10 and Figure 11. On 22 March, with an aim to curtail the spread of the novel coronavirus in the wake of four deaths and at least 39 infections, Bangladesh government deployed a nationwide lockdown effective from 26 March, 2020. Notice that, there are some jumps in the number of confirmed daily new infected cases data from 16 april to 20 april due to the increase of limited testing system (from 2000 samples to 2700 samples per day) Institute of Epidemiology, Disease Control and Research (IEDCR). Bangladesh is still struggling (around 6700 samples per day) Institute of Epidemiology, Disease Control and Research (IEDCR) to design a massive scale testing program as of May 11, 2020. Despite the fact, as we can see from Figure 10, our proposed model fits well for the historical real-time data. As time progresses, the estimated error declines and is hovering around 10% for the cumulative cases and daily new cases according to our calculated daily projected mean error. The estimated case-fatality rate in Bangladesh is at 2.7% which is kind of satisfactory, however without massive scale testing this rate could rise sharply incoming days. Moreover, the country’s estimated recovery rate is at 24% which again complements the real statistics precisely. Importantly, without an aggressive level mass-testing program it is impossible to portray the real outbreak scenario in Bangladesh. Table 6 illustrates the key features used to calibrate this scenario which are compatible with the previous clinical studies and relevant literature.

**Figure 10:**
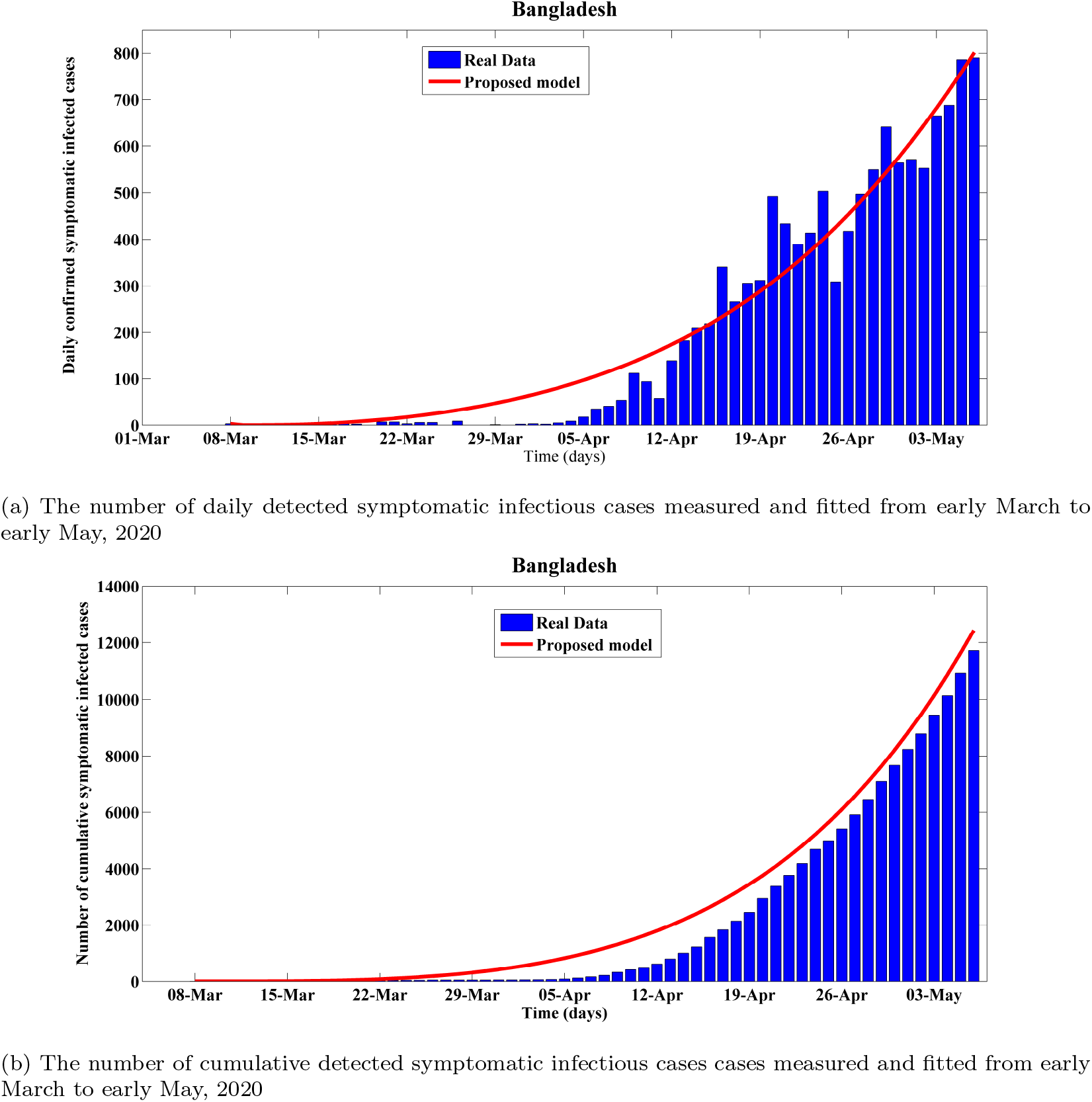
Fitting performance of calibrated *SEI_D_I_U_QHRD* model for Bangladesh from 8 March to early May, 2020

**Figure 11:**
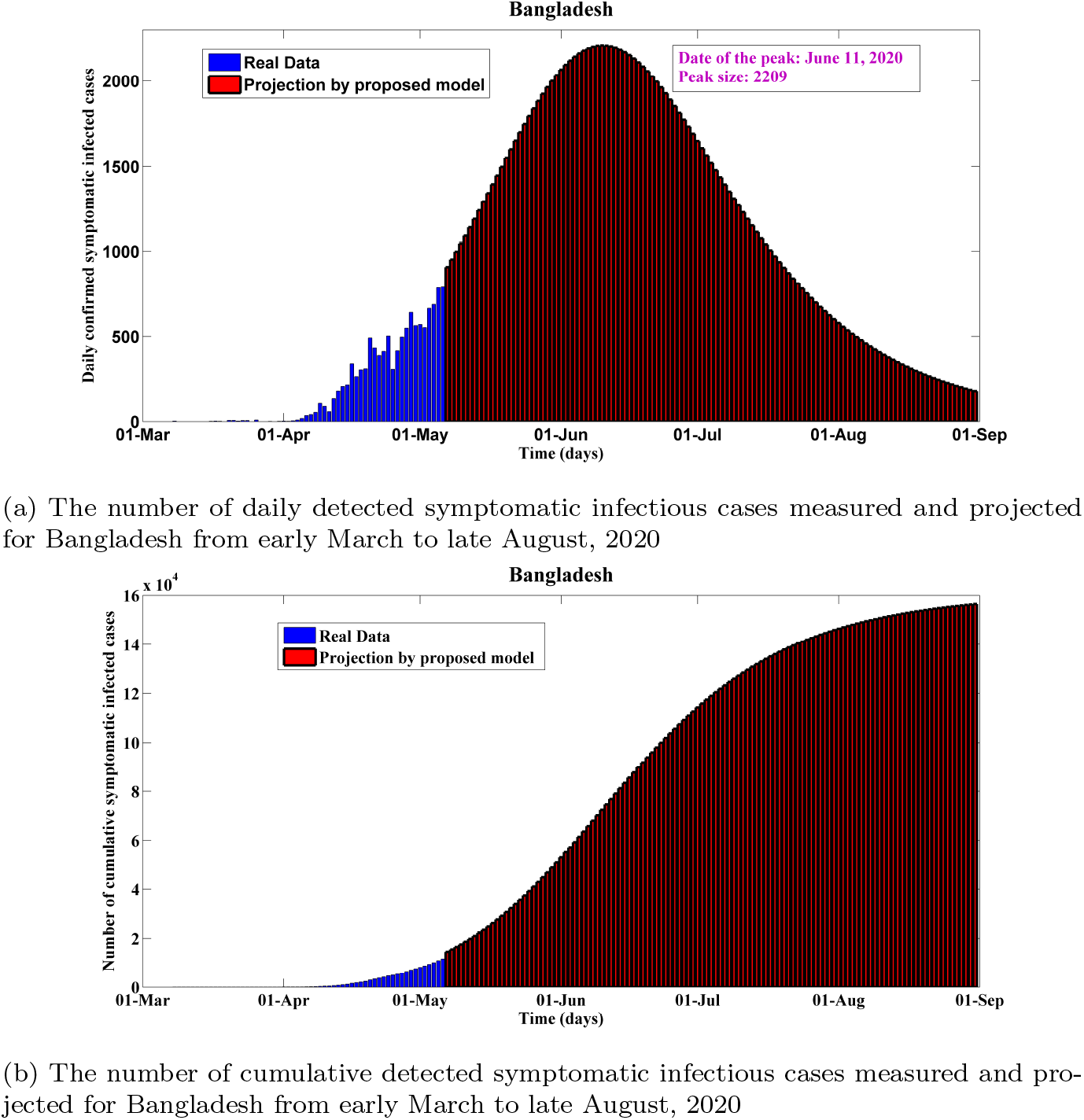
Predictions of the proposed SEIDIUQHRD model for Bangladesh from early March to late August, 2020

On 4 May, Bangladesh authorities intended to open up more factories, shopping malls and logistics operations, as they could diminish the economic impact of a coronavirus lockdown which they extended to May 16 Ministry of Health & Family Welfare of Bangladesh, (2020). This ease of lockdown could worsen the ongoing community transmission drastically. As we can see form Figure 11, the daily detected symptomatic infected cases (confirmed) will reach the peak (about 2209) around June 11, 2020 and then start to de-escalate. However, the probable peak time could occur in no time due to easing the coronavirus lockdown too quickly and this could bring a second wave of infections in the outbreak after post-peak period. The effective reproduction number is around 3.5 as of May 11, which again lies in prior established ranges Anastassopoulou et al., (2020); Liu et al., (2020). Our estimation is reasonably high which due to the fact that we have considered the infectiousness factor of the asymptomatic carriers. Massive level testing is highly required and recommended to identify the asymptomatic spreaders quickly. Unlike other mitigation strategies like reporting contacts, putting on face masks, and maintaining physical distancing, a massive test-and-isolate approach could control the disease burden of COVID-19. Otherwise, this basic reproduction number could increase upto 5.7 within 20 days and inhabitants of Bangladesh could see a disease catastrophe in near future. In near future, owing to various changing factors such as mitigation measures and mass people awareness such estimation and projection could differ significantly.

## 6. Global Sensitivity Analysis

PRCC analysis which is a global sensitivity analysis method that calculates the partial rank correlation coefficient for the model inputs (sampled by Latin hypercube sampling method) and outputs Marino et al., (2008); Blower et al., (1994); Nabi et al., (2020). The PRCC method assumes a monotonic relationship between the model input parameters and the model outputs.

The calculated PRCC values are between -1 and 1 and they are comparable among different model inputs. Quantitative relationship between the model input and model response can be determined by calculating the PRCC values. A positive PRCC value depicts that the model output can be increased by increasing the respective model input parameter or vice versa. In addition, a negative PRCC value indicates a negative correlation between the model input and output. The magnitude of the PRCC sensitivity measures the significance of the model input in contributing to the model output.

As our proposed epidemic model contains a moderate number of empirical parameters, uncertainty analysis can give considerable insights regarding the quantitative relationship between model responses and model input parameters. However, it is really challenging for complex models to determine the relationship with sufficient accuracy.

Importantly, we have got startling yet realistic results from our sensitivity analysis. As we can see from Figure 12, we found nearly the same qualitative and significant quantitative relationship between the number of symptomatic infectious individuals (one of the crucial model responses) and three parameters which are rate of getting quarantined of susceptible individuals (*q*), transition rate from exposed to infectious or quarantined (*κ*) which can also be referred as the inverse of the average incubation period of COVID-19 and recovery rate of undetected asymptomatic (undetected) infectious carriers for our proposed model.

In case of Russia, from Figure 12a recovery rate of undetected asymptomatic carriers (*ϕ_U_*) is the most negatively influential parameter when the number of detected infectious individuals is our selected model response. The PRCC index is found to be −0.585. In addition, rate of entering into home-quarantine or self-quarantine of susceptible individuals (*q*) and the fraction of wrongly quarantined people who become susceptible after certain latent period (*r*_1_) are the other two influential empirical features with PRCC indexes are −0.5672 and 0.567 respectively.

In case of Brazil, rate of getting home-quarantined or self-quarantined of susceptible individuals (*q*), the fraction of quarantined people who become susceptible due to avoiding home-quarantine (*r*_1_), and the inverse of the COVID-19 mean latent period are the most influential parameter on the symptomatic infectious population size (*I_D_*(*t*)). The corresponding PRCC indices are −0.62, 0.53 and −0.499. The figure 12b qualitatively elucidates that a high quarantine rate of the susceptible individuals can curtail the number of symptomatic infected individuals. In broader view, high efficacy of home or self-quarantine and a lower unlockdown rate could flatten the *I_D_*(*t*) curve in Brazil.

In case of the India, rate of getting home-quarantined or self-quarantined of susceptible individuals (*q*) and the inverse of the COVID-19 incubation period are the most influential parameter on the symptomatic infectious population size (*I_D_*(*t*)) and the corresponding PRCC indexes are −0.64 and −0.62. Both the parameters are negatively sensitive to the size of detected infected individuals which is illustrated in the figure 12c. This elucidates that a higher quarantine rate of the susceptible individuals can reduce the number of symptomatic infected individuals. Therefore, it is obvious that early unlockdown in India could worsen the outbreak situation in the blink of an eye.

**Figure 12:**
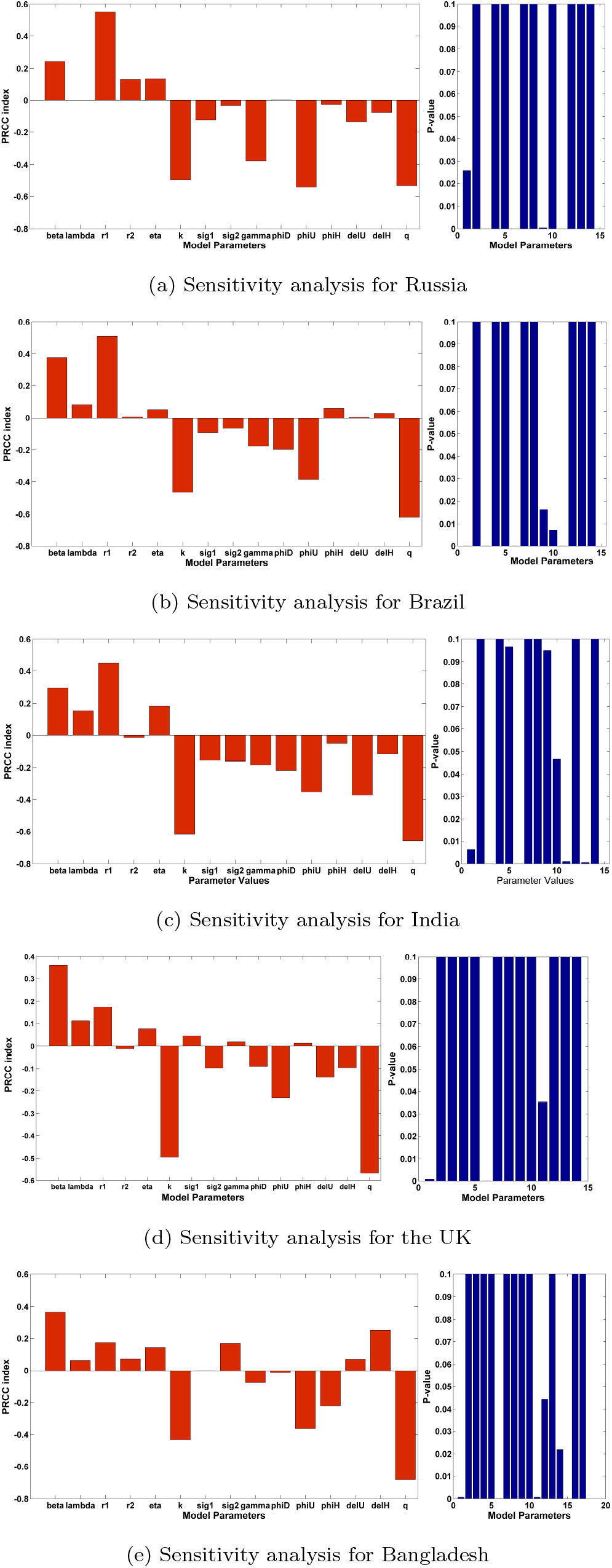
Sensitivity of the symptomatic infected cases while changing parameters in the proposed *sei_d_i_u_qrhd* model as indicated by the PRCC index for five different countries

In case of the UK, rate of getting home-quarantined or self-quarantined of susceptible individuals (q) and the inverse of the COVID-19 incubation period are the most influential parameter on the symptomatic infectious population size (*I_D_*(*t*)) and the corresponding PRCC indices are −0.58 and −0.499. Both the parameters are negatively sensitive to the size of detected infected individuals which is illustrated in the figure 12d. This elucidates that a higher quarantine rate of the susceptible individuals can curtail the number of symptomatic infected individuals. Nevertheless, early easing lockdown measures could bring a second wave of infection in the UK in no time.

For Bangladesh, it has been found in our analysis, home-quarantine rate is the most negatively sensitive parameter on the size of symptomatic infectious individuals and the corresponding PRCC index is −0.68 which is the highest among our five studied cases. The result in Figure 12e elucidates that if the people in Bangladesh start breaking social distancing restrictions extensively, then it would be difficult to control the disease outbreak there. As of May 11, when Bangladesh is seeing continuous spikes in COVID-19 infections every day, the government has announced an easing of movement for all on a limited scale with a view to reviving the country’s economy. Moreover, the length of the inverse of the latent period is the another negatively sensitive parameter with PRCC index lying at −0.43.

## 7. Concluding Remarks

We have proposed a methodology for the calibration of the key epidemiological parameters as well as forecasting of the outbreak dynamics of COVID-19 pandemic in Russia, Brazil, India, Bangladesh and the UK, while considering publicly available data from late January 2020 to early May 2020 with an introduction of an real-time differential *SEI_D_I_U_QHRD* epidemic model which can give more accurate, realistic and precise short-term predictions. Baseline parameter ranges are chosen from the recent clinical studies and relevant literature concerning the COVID-19 infection. Trust-region-reflective algorithm which is one of the robust least-squares fitting techniques, has been deployed to calibrate the proposed model parameters. Numerical results on the recent COVID-19 data from Russia, Brazil, India, Bangladesh and the UK have been analyzed with an aim to determine probable peak dates and sizes for the above mentioned countries. Based on the projection as of May 11, 2020, Russia will reach the peak in terms of daily infected cases and death cases around end of May. Brazil and India will reach the peak in terms of daily symptomatic infectious cases and death cases around beginning of June. In addition, the United Kingdom might face a second wave of infection provided that lockdown is lifted quickly. Global sensitivity analysis results depict that home-quarantine or self-quarantine is the most effective measure for controlling the transmission and spread of the novel coronavirus infection. To fade out the pandemic, it is compulsory to identify potential carriers and asymptomatic infectious spreaders through and massive scale testing scheme and without sufficient level of evidence of controlled low transmission rate of COVID-19 in any certain territory, we cannot afford lifting up physical distancing measures.

## Data Availability

All data is available.

## Notes

### Competing Interest Statement

The authors have declared no competing interest.

### Funding Statement

No funding is involved.

